# Red flags for potential serious pathologies masquerading as musculoskeletal neck pain: a protocol for a systematic review of clinical practice guidelines

**DOI:** 10.1101/2023.06.20.23291691

**Authors:** Daniel Feller, Alessandro Chiarotto, Bart Koes, Filippo Maselli, Firas Mourad

## Abstract

**Background:** Although benign most of the time, neck pain (NP) has been observed as the early manifestation of various serious cervical pathologies (e.g., malignancies, fractures). Red flags (RF) are signs and symptoms that raise suspicion of serious spinal pathology. Considering that it has been estimated that the incidence of delayed diagnosis of serious cervical pathologies ranges from 5% to 20%, investigating RFs for NP remains a priority for an informed practice and the patient’s safety. Therefore, the following systematic review will aim to identify red flags recommendations to triage serious pathologies in current clinical practice guidelines (CPGs) for patients with NP, to evaluate the consistency between different CPGs regarding RF recommendations, and to investigate on what study type the recommendation of CPGs are based.

**Material and methods:** We will search for CPGs for patients with specific or non – specific NP by searching MEDLINE (via PubMed), EMBASE, and PEDro electronic databases. Guidelines will also be searched through forward and backward citation tracking strategies (Web of Science), by consulting experts in the field, and by checking guideline organization databases. Also, we will screen the references of two recently published systematic reviews on CPGs for NP. For all the CPGs included, we will extract bibliographic information, the serious pathologies considered, the RF considered for all serious pathologies, the reference used to support all the RF cited in the guideline, and, if available, the diagnostic accuracy data for all the RF. Two authors will independently perform the study selection and data extraction processes.

**Data synthesis:** Results will be presented descriptively and using graphs and tables. We will evaluate the consistency among the guidelines in their endorsement of red flags using Fleiss’ kappa. Fleiss’ kappa will be presented separately for all serious pathologies the CPGs consider.

## BACKGROUND

Neck pain (NP) is a complex biopsychosocial disorder associated with problematic physical and psychological symptoms and has been estimated as the fourth global contributor to Years Lived with Disability. ^1–3^ Although benign most of the time, NP has been observed as the early manifestation of various serious cervical pathologies (namely, spinal masqueraders), making their diagnosis a real challenge for clinicians. ^4,5^ it has been estimated that the incidence of delayed diagnosis of serious cervical pathologies ranges from 5% to 20% with potentially life-threatening consequences. ^6,7^ Red flags (RF) are signs and symptoms that raise suspicion of serious spinal pathology. Notably, there is a paucity of literature on RFs for NP; that is, RFs are typically informed by the LBP literature. ^1^ Although the recent release of the International Federation of Orthopaedic Manipulative Physical Therapists Cervical Framework, differential diagnosis of the cervical spine is a critical topic. ^5,10^ Therefore, investigating RFs for NP remains a priority for an informed practice and the patient’s safety.

Against this background, with the present systematic review, we will aim:

1. To identify red flags recommendations to triage serious pathologies in current clinical practice guidelines (CPGs) for patients with NP
2. To evaluate the consistency between different CPGs regarding RF recommendations
3. To investigate on what study type the recommendation of CPGs are based (e.g., a systematic review of primary diagnostic studies, case reports)

## MATERIAL AND METHODS

### Inclusion criteria

We will include CPGs, defined by the World Health Organization as “systematically developed evidence-based statements that assist providers, patients, policymakers and other stakeholders to make informed decisions on health care and public health policy”. ^11^ A document will be regarded as a CPG if it respects the following criteria (adapted from the PEDro criteria for evidence-based clinical practice guidelines ^12^):

1. Produced under the auspices of a health professional association or society, public or private organisation, health care organisation or plan, or government agency
2. A systematic literature search and review of existing scientific evidence was performed during the guideline development, or the guidelines were based on published systematic reviews
3. The clinical practice guideline contains systematically developed statements that include recommendations, strategies, or information to guide decisions about appropriate healthcare

We will include CPGs focusing on specific and non-specific NP. Specific NP (or NAD III) refers to pain and symptoms directly attributed to a biological (non-serious pathology or disease) process associated with the musculoskeletal system (e.g., cervical radiculopathy). Non-specific NP (or NAD I/II) refers to musculoskeletal pain disorders and associated impairments (e.g., cervicogenic headache, whiplash). Non-specific NP is determined by no underlying pathology (i.e., specific features or pain generators are not identifiable). We will not include Serious NP (or NAD IV) which refers to a serious pathology or conditions mimicking a musculoskeletal NP that warrants further investigation and referral to the appropriate healthcare professional. Serious NP are identifiable during the history/examination by the presence of risk factors and clinical features (namely, RFs) potentially associated with serious pathologies, including malignancy, inflammatory disorders, fracture, and infection. ^1^

We will not apply any restrictions regarding publication date and language. Non-English and non-Italian guidelines will be translated using “DeepL Translate” (https://www.deepl.com/). Subsequently, we will ask a native speaker to verify the translation accuracy regarding the section on red flags.

We will exclude CPGs not specifically focused on NP, such as guidelines in which NP is only briefly mentioned in the context of other disorders or a more complex topic (e.g., management of chronic pain in general). In addition, if multiple versions of the same CPG from a certain professional body are present, we will only include the most up-to-date version.

### Study selection process

We will search for CPGs for patients with NP by searching MEDLINE (via PubMed), EMBASE, and PEDro electronic databases.

We will search PubMed using the following search string:

*(“Neck Pain”[MeSH Terms] OR “Neck Pain”[Title/Abstract] OR “cervical pain”[Title/Abstract] OR “neckache*”[Title/Abstract] OR “neck ache*”[Title/Abstract] OR “cervicodynia”[Title/Abstract] OR “cervical vertebrae”[Title/Abstract] OR “cervical spine”[Title/Abstract] OR “cervico*”[Title/Abstract] OR whiplash[Title/Abstract] OR “Radiculopathy”[Mesh] OR “Cervical radiculophaty”[Title/Abstract] OR “Post-Traumatic Headache”[Mesh]) AND (“guideline*”[Title/Abstract] OR “Guideline” [PT] OR “Guidelines as Topic”[Majr] OR Consensus Development Conference [PT])*

EMBASE will be searched using the following search string:

*(‘Neck Pain’/exp OR ‘Neck Pain’:ti,ab OR ‘cervical pain’:ti,ab OR neckache*:ti,ab OR ‘neck ache*’:ti,ab OR cervicodynia:ti,ab OR ‘cervical vertebrae’:ti,ab OR ‘cervical spine’:ti,ab OR cervico*:ti,ab OR whiplash:ti,ab OR Radiculopathy/exp OR ‘Cervical radiculophaty’:ti,ab OR ‘Post-Traumatic Headache’/exp) AND (‘guideline*’:ti OR ((practice* OR clinic*) NEAR/2 guideline*):ti,ab OR ‘practice guideline’/exp OR ‘consensus development’/exp)*

Guidelines will also be searched through forward and backward citation tracking strategies (Web of Science), by consulting experts in the field (top ten experts on neck pain according to ExpertScape), and by checking guideline organization databases. Also, we will screen the references of two recently published systematic reviews on CPGs for NP. ^13,14^

The guideline organization databases that we will search will be the following: the “Canadian Medical Association infobase of clinical practice guidelines” (https://joulecma.ca/cpg/homepage), the “Istituto Superiore Sanità - Sistema Nazionale LineeGuida” (https://snlg.iss.it), the “Guidelines International Network” (https://guidelines.ebmportal.com/), the “National Institute for Clinical Excellence – NICE” (https://www.nice.org.uk/), the “OPTIMa collaboration” (https://idrr.ontariotechu.ca/publications), the “Guideline Central” (https://www.guidelinecentral.com/), and the “Scottish Intercollegiate Guidelines Network – SIGN” (https://www.sign.ac.uk/our-guidelines/).

Duplicates will be eliminated using the Deduplicator function of “Systematic Review Accelerator”.^15^

Two researchers will independently perform the study selection process by title/abstract and then by full text. Any disagreement will be resolved by consensus or by the decision of a third author. We will use the online electronic systematic review software package (Rayyan QCRI) to organize and track the selection process. ^16^

### Data extraction

Two reviewers will be informed and trained on eligibility criteria, and conduct the data extraction process independently using a standardized form. Any discrepancies will be resolved with a consensus between the two authors and eventually by a third author’s decision.

We will extract the following data from all the CPGs included:

1. publication year,
2. language,
3. association(s) or society(ies) producing the guideline,
4. serious pathologies considered (e.g., malignancy, fracture, infection, congenital craniovertebral anomalies, cervical arteries dysfunctions),
5. RF considered for all serious pathologies, and if these RF are presented in general or subdivided for single pathologies,
6. reference used to support all the RF cited in the guideline, or, in the alternative, if the RF was supported by the consensus of the guideline committee only, or if no information was given to support the endorsement of the RF, and
7. if available, diagnostic accuracy data for all the RF

### Data synthesis

Results for the first and third review questions will be presented descriptively, and using graphs and tables.

For the second review question, we will evaluate the consistency among the guidelines in their endorsement of red flags using Fleiss’ kappa. ^17^

Fleiss’ kappa will be presented separately for all serious pathologies the CPGs consider.

## Data Availability

All data produced in the present study are available upon reasonable request to the authors

## ETHICS AND DISSEMINATION

After completion of the study, a manuscript with results will be submitted for publication in a peer-reviewed journal.

## AUTHOR CONTRIBUTIONS

D.F., F.Mo., and A.C. conceived and designed the study protocol. All the authors provided input and approved the final version of the protocol.

